# Patterns of self-medication among university students in San Jose, Costa Rica

**DOI:** 10.1101/19012047

**Authors:** Maria Natalia Norori

## Abstract

**Background:** Self-medication is defined as the use of drugs to treat self-diagnosed symptoms without the supervision of healthcare physicians. Self-medication is a growing public health phenomenon and is associated with risks such as misdiagnosis and drug toxicity. This study aimed to identify the patterns associated with the practice of self-medication among university students in San José, Costa Rica.

**Methods:** A descriptive cross-sectional study was designed and conducted to identify variables associated. Information was collected on the conditions treated, medications used and their sources.

**Results:** The study found that self-medication is highly prevalent among Costa Rican university students. 91.4% of the sample reported self-medicating, with each student consuming an average of 2.15, ± 1,08 drugs. The most Frequently used active ingredients were paracetamol and Ibuprofen. Results also show a relation between the most consumed types of drugs and the principal causes of drug intoxication reported by Costa Rica’s National Poison Center. 77.8% of the participants considered self-medication a risky practice.

**Conclusions:** Self-medication is common among Costa Rican university students. The prevalence found is higher than that reported in previous studies conducted in the country. These findings suggest the need to implement prevention campaigns and regulatory policies to ensure the safe consumption of medical drugs.

## Background

The World Health Organization defines self-medication as the use of drugs to treat self-diagnosed disorders or symptoms, or the intermittent or continued use of a prescribed drug for chronic or recurrent diseases or symptoms without the supervision of a medical physician (1). Self-medication is an important public health phenomenon; which may delay diagnosis and facilitate the emergence of resistant diseases and microorganisms, affecting quality of life and global health as a whole (2).

Previous studies have shown that self-medication is associated with risks such as excessive drug dosage and use, drug interactions and polypharmacy, incorrect self-diagnosis and misdiagnosis, poor adherence and delay in medical treatment (2,3). Nonetheless, responsible self-medication has also shown multiple benefits, including a reduction of the burden of health services, economic improvement, a reduction of waiting lists and an increase in patients’ active role in their health (3).

In Costa Rica, there are about 180 drugs sold over-the-counter in supermarkets, pharmacies and other establishments (4). Self-medication is a practice commonly carried out by the Costa Rican population. Past studies show that 81% of the population actively self-medicates (4). In most cases, individuals start self-medicating as teenagers and young adults (5). Almost half of the population that practices self-medication has reported the easy identification of the symptoms and pathologies treated as one of the main reasons for doing so. (4, 5).

Although self-medication is an alarming phenomenon and studies have been conducted to measure its prevalence in the country (4,5), there are no recent studies that focus on measuring the prevalence of self-medication among Costa Rican university students. Studies conducted in the university population of different countries show that the prevalence of self-medication is higher in university students than that in the rest of the population (6, 7).

Having up-to-date evidence-based data on drug consumption patterns is essential for the implementation of policies and regulatory strategies that promote rational drug use (8). The objective of this study is to identify the prevalence and patterns associated with the practice of self-medication among university students in San José, Costa Rica.

## Methods

A descriptive cross-sectional study was conducted between June 2019 and August 2019 with students from a university located in San Jose, Costa Rica. Participants were selected by random selection and had to be enrolled in a university to be eligible for the study.

To estimate the prevalence and self-medication patterns in the population, a 10-question questionnaire was designed based on previous studies of a similar approach (8) (11) and reviewed by three public health professionals. A pilot study was conducted in June 2019 with 20 students from all fields to test the instrument and assess its understanding and effectiveness. The questionnaire was adjusted as per feedback from the piloted sample, and data of the pilot study was not included in the results.

Data concerning sociodemographic and general characteristics, medication consumption habits, prevalence and frequency of medication use, forms of acquisition, sources of advice, frequent symptoms prompting self-medication and pharmacological drugs used. Prior to answering the questionnaire, all participants were given a brief explanation of the intention of the study and gave their informed consent.

To calculate the sample size, a 95% confidence interval and 0.05 significance level were used. The calculated sample size was increased to 15% to compensate for possible losses and memory bias, thus obtaining an adjusted sample size of 157.

Descriptive statistics such as frequency, mean, and standard deviation were used to present participants and their characteristics. Data analysis was carried out using the SPSS software version 25.0 at a level of significance of P<0.05.

## Results

A total of 139 students answered the questionnaire correctly and were included in the data analysis. Of the 139 students included in the study, 56.5% were female and 70% did not work in any company or business of their own. 49.6% of the students were aged between 21-25 years old, while 39.6% were aged between 16 and 20 years old. Only 10.6% of the sample were between 26 and 30 years of age.

The university where the sample was collected has 5 faculties: Faculty of Health Sciences, Faculty of Engineering and ICT, Faculty of Business Sciences, Faculty of Social Sciences and Faculty of Art and Design. 36.7% of the participants were students of the Faculty of Health Sciences, while 34.5% studied in the Faculty of Engineering and ICT. 12.9% and 11.5% of the sample were students of the Faculty of Business and Social Sciences, respectively. Only 4.6% of the students were enrolled in the Faculty of Arts and Design. The general characteristics of the participants are described in Table 1.

**Table 1:** General characteristics of the sample

| Variable | sample (%) | Prevalence of self-medication(%) |
| --- | --- | --- |
| **Sex** |  |  |
| Male | 59 (42.4) | 53 (89.8) |
| Female | 80 (57.6) | 74 (92.5) |
| **Age range** |  |  |
| 16-20 | 55 (39.6) | 47 (85.5) |
| 21-25 | 69 (49.6) | 66 (95.7) |
| 26-30 | 15 (10.8) | 14 (93.3) |
| **Faculty** |  |  |
| Business Sciences | 18 (11.9) | 15 (83.3) |
| Social Sciences | 16 (12.5) | 16 (100) |
| Health Sciences | 51 (36.7) | 46 (90.2) |
| Engineering and ICT | 48 (34.5) | 44 (91.7) |
| Arts and Design | 6 (4.3) | 6 (100) |

Self-medication was reported by 91.4% (n=127) of participants. When asked about their frequency of consumption, 68.3% of the sample reported self-medicating once a month, although 23% reported practicing self-medication every 15 days. Out of the percentage of the sample that reported self-medicating, 58.3% were female and the remaining were male. Students enrolled in the Faculty of Business Sciences reported self-medicating the least, with a prevalence of self-medication of 83.3%, followed by the students of Health Sciences, who reported a 90.2%prevalence. On the other hand, 100% of the students enrolled in the Faculty of Social Sciences reported practicing self-medication.

The principal symptoms associated with self-medication were headache (75.4%), flu (63.5%) and allergies (34.9%). Acid reflux, stomach ache, diarrhea, and infectious diseases were also reported on lower percentages (Figure 1).

Each student reported consuming an average of 2.15, ± 1.08 drugs. As for the types of drugs consumed, participants predominantly reported using paracetamol (53.5%) and other NSAIDS (45.7%), followed by antihistamines (15%), anti-flu decongestants (15%) and antacids (15%). 12.6% of the sample does not remember the name of the medication they consumed, but do recall self-medicating. A smaller percentage of the sample reported self-medicating with antibiotics, contraceptives, and antiemetics.

62.4% of the students that reported self-medicating don’t believe their symptoms require medical attention, and 40.8% reported not having time to consult a physician. Interestingly, only 16% of participants listed a lack of economic resources as a reason to self-medicate. (Table 2).

**Table 2:** Reasons for self-medication among Costa Rican university students

| Reasons associated | Number of cases (%) |
| --- | --- |
| Does not consider it necessary to see a doctor | 78 (62.4) |
| Lack of time | 51 (40.8) |
| Lack of economic resources | 20 (16) |
| Lack of medical insurance | 11 (8.8) |
| Other | 13 (10.4) |

The primary sources of advice for taking medications for all students were family members (72.6%) and pharmacy staff (37.9%). Other primary sources reported are shown in Figure 2.

Only 44.9% of the students reported reading the instructions for use. 85% reported having a home pharmacy. Logistic regression showed a significant association between self-medication and having a home pharmacy.

Despite the high prevalence of self-medication among the Costa Rican student population, 77.8% of the sample considered self-medication a risky practice.

## Discussion

In the present study, 91.4% of the participants reported self-medicating. This result is similar to that reported in similar studies conducted with university students in Spain (90%) (6), Slovenia (96%) (9) Peru (98%) (10), United States (89%) (11) and Mexico (96%) (12). Interestingly, the prevalence of self-medication is higher than that obtained by the survey “Actualidades 2017”, conducted by the School of Statistics of the University of Costa Rica, which estimated that 81.7% of Costa Ricans self-medicate (13).

The high prevalence obtained can be attributed to different factors, such as easy accessibility to over-the-counter medications, and the fact that 85% of respondents have a home pharmacy. It has been proven that having medications stored in the place of residence increases the risk of self-medication (13). This confirms the important role that the pharmaceutical industry plays in controlling such practice.

There were no significant differences in the prevalence of self-medication between healthcare and non-healthcare students (p= 0.456), which differs from that found in studies in Slovenia (9), Mozambique (14), which reported a lower prevalence of self-medication in healthcare students, and resembles what was reported by Spanish researchers (6).

As for the types of drugs, analgesics were the group of drugs most consumed by self-medication. More than half of the sample reported self-medicating with paracetamol (53.5%), and 45.7% reported consuming other NSAIDs. The use of analgesics and anti-inflammatories is related to the main symptoms associated with self-medication: Headache (75.4%) and flu (63.5%). These findings are similar to those reported in similar studies (6, 9-11). Although analgesics are extremely useful drugs, the non-responsible consumption of paracetamol and NSAIDs is related to hepatotoxicity, nephrotoxicity, and ulceration of the gastric mucosa (15). According to records issued by the National Poison Center of the Costa Rican social security system, Caja Costarricense de Seguro Social, drug intoxication causes 58% of poisonings in Costa Rica. In 2014, a daily average of 12 cases of drug poisoning was recorded. The majority of cases reported were caused by intoxication with paracetamol, which was also the drug most of this study’s participants reported self-medicating with. (16). The implementation of regulatory policies is important to regulate its consumption and minimize its risks.

It should be noted that the present study found a higher consumption of antihistamines than that reported in similar studies conducted in other countries (11,12,14). This may be related to the prevalence of rhinitis and allergies in the Costa Rican population (42.6%), as well as environmental and geographical factors (17). In Costa Rica, the medications with the highest prevalence of poisoning are paracetamol, antihistamines, and ibuprofen (16,18). These drugs correspond to those most consumed by the sample in the study, which suggests there is an urgent need to deepen existing efforts to inform and educate about the safe use of medications.

The fact that the main reasons for self-medication are lack of time and the lack of necessity to visit a physician confirm the role that access to information through the internet plays to make users feel aware of pathologies and symptoms. This is a double-edged sword, as it increases the role that consumers play in their healthcare decisions, but can also lead to misdiagnoses and poor adherence to treatments (19). Waiting lists in social security facilities and the high costs of private medical consultants also influence this practice (20).

75% of the sample listed family members as main sources of information for the medicines they consumed, which leads one to think self-medication is an accepted practice among Costa Ricans. It is important to carry out complementary studies in other population groups in order to establish possible intervention points to promote responsible self-medication and avoid the risks that come with the practice.

This study has limitations that need to be addressed, among which are the small sample size used, the fact that the sample refers to a specific geographic area, the use of a questionnaire as the data collection instrument, and possible memory bias. Despite its limitation, the present study offers the possibility to guide other researchers and health organizations working to address the issue.

## Conclusion

The consumption of over-the-counter drugs among Costa Rican university students deserves further study. The high prevalence of self-medication found demonstrates the need for local health authorities to implement regulatory and managerial strategies to control the practice, and prevention campaigns to educate the population and promote responsible self-medication and rational drug consumption. Improved knowledge and understanding might help increase the benefits and reduce the risks associated with self-medication.

## Data Availability

Data has been deposited under a cc by 4.0 license in https://zenodo.org/record/3539589#.XcsQuEVKhZ0

## Abbreviations

NSAID: Nonsteroidal anti-inflammatory drug

## Declarations

### Ethics approval and consent to participate

This study was approved by the ethics committee of Universidad Latina de Costa Rica. Verbal consent was obtained after explanation of the objectives of the study from all participants, no personal information is provided and data was collected anonymously and verbal consent was approved by the ethics committee.

### Consent for Publication

Not applicable

### Availability of data and materials

The datasets used and/or analyzed during the current study are available from the corresponding author on reasonable request.

### Author contributions

The corresponding author was responsible for the design of the work, acquisition, analysis, and interpretation of data, the draft, revision and approval of the submitted manuscript. There were no earlier authors involved or removed from the study.

## Acknowledgements

Not applicable

